# Estimating COVID-19 Antibody Seroprevalence in Santa Clara County, California. A re-analysis of Bendavid et al.

**DOI:** 10.1101/2020.04.24.20078824

**Authors:** Stephen T. Bennett, Mark Steyvers

## Abstract

A recent study by Bendavid et al. claimed that the rate of infection of COVID-19 in Santa Clara county was between 2.49% and 4.16%, 50-85 times higher than the number of officially confirmed cases. The statistical methodology used in that study overestimates of rate of infection given the available data. We jointly estimate the sensitivity and specificity of the test kit along with rate of infection with a simple Bayesian model, arriving at lower estimates of the rate of COVID-19 in Santa Clara county. Re-analyzing their data, we find that the rate of infection was likely between 0.27% and 3.21%.

## Introduction

A recent Stanford study by Bendavid et al. [1] found that 50 of 3330 people living in Santa Clara county tested positive for the novel Coronavirus, COVID-19. The authors of the study claim that this rate of positive test results, combined with the specificity and sensitivity of the test kit they used, were sufficient to believe that 2.49% to 4.16% of the people living in Santa Clara county were infected with COVID-19 as of April 4th, 2020. This study was quick to make headlines [2] [3] [4], but those estimates are too high given the data available.

The specificity of the test kit is too low to come to such confidently high estimates based on their sample. The authors give confidence intervals for the specificity of the test kit that ranged from 98.1% to 100%. If we take this range seriously, then it would be possible to explain their results even if no one in Santa Clara were sick. Indeed, a sample of 3330 healthy people tested by a test kit with 98.1% specificity would have 63.27 false positives on average, more than the positive test results found by Bendavid et al.

In this paper, we provide the results of a simple Bayesian model in which the specificity and sensitivity of the test kit are jointly inferred along with the rate of infection in Santa Clara at large. This model allows us to correctly propagate uncertainty about the accuracy of the test kit when estimating the rate of infection. Under our model, we infer the posterior density of the rate of infection, specificity, and sensitivity and compute 95% Highest Density Intervals for each of them to summarize our findings.

We focus solely on the statistical methodology used to infer the rate of infection from the sample and make no claims about the appropriateness of the other methods used in the Bendavid et al. study, such as demographic re-weighting or the sampling procedure.

## Methods

### Data

In order to inform the specificity and sensitivity of the test kit, we assume “Scenario 3” as described by the authors. That is, we assume that both their data and the data provided by the manufacturer are useful for estimating the specificity and sensitivity of the test kit used on their Santa Clara sample. When the manufacturer assessed their test kit, it was able to correctly identify 369 of 371 pre-COVID samples known to be uninfected. The authors tested an additional 30 pre-COVID samples from hip surgery, all of which the test kit correctly identified. Combining both sources, the test kit falsely identified 2 out of 401 samples that could not have had the disease. To inform its ability to detect the disease in infected samples, the manufacturer found the test kit correctly identified 153 of 160 samples from clinically confirmed COVID-19 patients. The authors tested samples from an additional 37 clinically confirmed patients, 25 of which were correctly identified. In total, the test kit correctly identified 178 of 197 presumed-infected individuals.

The authors collected 3330 volunteers from Santa Clara county recruited via Facebook and tested them with this test kit. 50 of these people tested positive for COVID-19.

### Priors

We assume a-priori that sensitivity, specificity, and the rate of infection are each distributed according to a Beta(2,2) distribution. The probability density function associated of this distribution is in panel 1 of Figure 2. This prior is uninformative, reflecting the belief that these variables could take any of a wide range of possible values. The appendix analyzes the sensitivity of our conclusions based on these priors.

**Figure 1:**
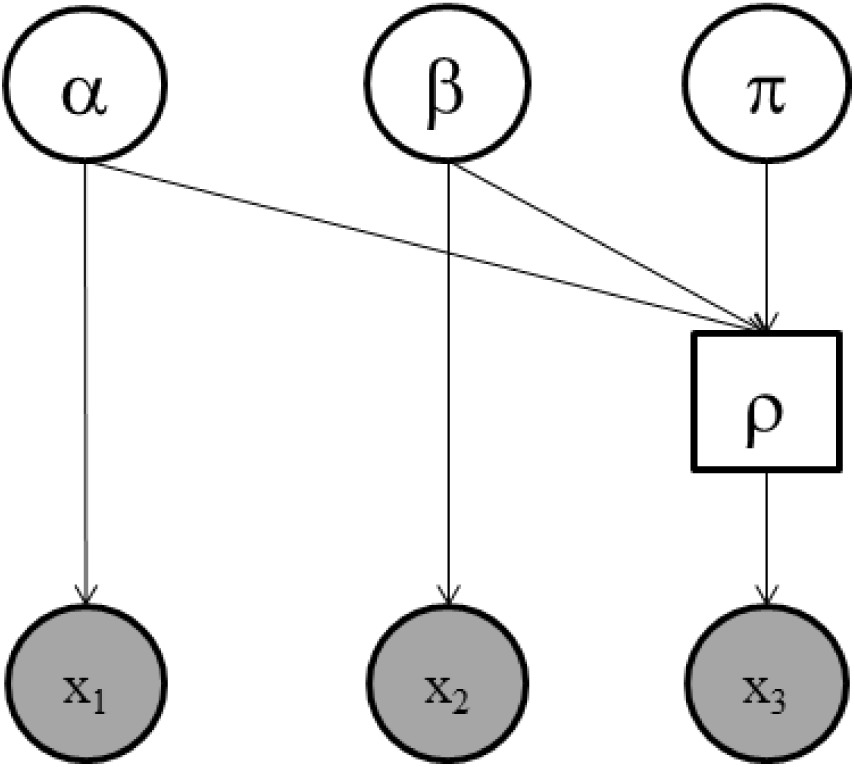
Graphical representation of our model.

**Figure 2:**
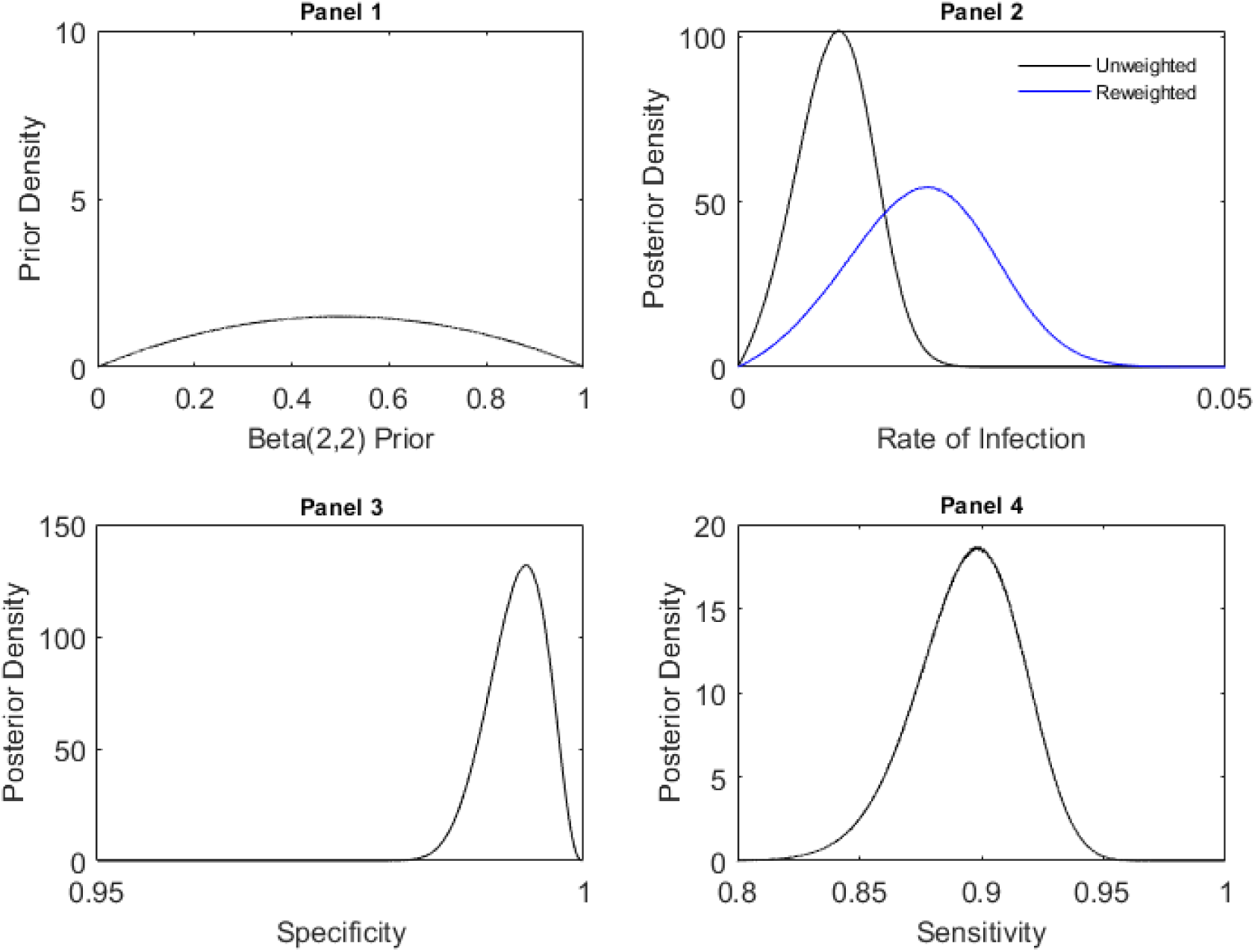
**Panel 1**: Prior distribution for Specificity, Sensitivity, and the Rate of Infection. **Panel 2**: Posterior distribution of the rate of infection in Santa Clara, before and after re-weighting based on demographic data. **Panel 3**: Posterior distribution of the specificity of the test kit. **Panel 4**: Posterior distribution of the sensitivity of the test kit.

### Model Specification

We assume a simple Bayesian model that explains the test results given the uncertain specificity (*α*), sensitivity (*β*), and rate of infection in Santa Clara (*π*). From these values, we compute the rate of positive test results (*ρ*). The Santa Clara sample (*x*3) is distributed according to a Binomial distribution given *ρ*. The test kit data (*x*1 & *x*2) are distributed Binomially with rate parameters based on the sensitivity and specificity of the test kit. A simple graphical representation is shown in Figure 1. Circles that are shaded gray are our observed data. Circles that are not shaded we infer by sampling. This is done jointly for all variables and all data.

### Sampling Details and Code

We use JAGS, a Markov chain Monte Carlo (MCMC) sampler, to condition the model upon the data. We collected 5,000,000 samples of the posterior distribution from 7 chains after 1,000,000 burn-in samples.

All data and code associated with the project is available via the Open Science Framework: https://osf.io/5qhgb/.

### Demographic re-weighting

Bendavid et al. re-weight the estimate of the rate of infection based on the demographic data of their sample. This factor was intended to compensate for the non-representative sample they recruited for their study. They propagate uncertainty from the inferred number of actual cases in their sample to the inferred number of actual cases in the population at large by multiplying the sample’s rate of infection by 1.87. We make no claim about the suitability of this re-weighting, but include it in order to match our results with those of the original authors. Results are presented both with and without this factor.

## Results

### Specificity and sensitivity

The posterior distributions for specificity and sensitivity are shown in panels 3 and 4 of figure 2. There is substantial uncertainty about both of these values based on the data available. The 95% highest density interval for the specificity of the test kit is [98.73%, 99.83%] and the 95% highest density interval for the sensitivity of the test kit is [85.11%, 93.56%]. If this lower bound of specificity (98.72%) were true, in expectation we would observe 42.6 positive test results out of the 3330 individuals tested even if no one in Santa Clara were infected with COVID-19.

### Rate of Infection

The posterior distribution of the rate of sick individuals in Santa Clara county is shown in panel 4 of Figure 2. The 95% highest density interval associated with this posterior distribution is [0.27%, 1.72%]. Replicating the demographic re-weighting done by the original authors, this interval becomes [0.49%, 3.21%].

## Discussion

Reanalyzing the data from Bendavid et al., we find that the rate of infection was likely between 0.27% and 3.21% in early April. This interval is substantially lower than the interval used to draw conclusions by the original authors (2.49% to 4.16%). Based on our analyses, there is a 79.47% chance that the true rate of infection was below the lower bound of this interval even after re-weighting based on demographics.

We re-analyze some of the conclusions of the original authors based on our estimate of the rate of infection, 0.27% to 3.21%. This rate of infection means between 5,000 and 65,000 people were infected in Santa Clara county. As of April 1st, 956 cases had been confirmed in Santa Clara. This would correspond to between a 5-to-1 and 65-to-1 ratio of observed-to-total cases of COVID-19 (the underascertainment rate).

## Acknowledgments

We want to thank the Bendavid et al. Without them we would not have the data to do this analysis and be forced to resort to very speculative priors about the rate of infection in Santa Clara county.

## Appendix

### Sensitivity Analysis

We analyzed how our results differed as a function of the priors used in the model. Table 1 shows the results assuming all distributions have prior Beta(x,x) for various values of x. These results are each based on 500,000 samples from JAGS after 100,000 burn-in samples.

**Table 1:**
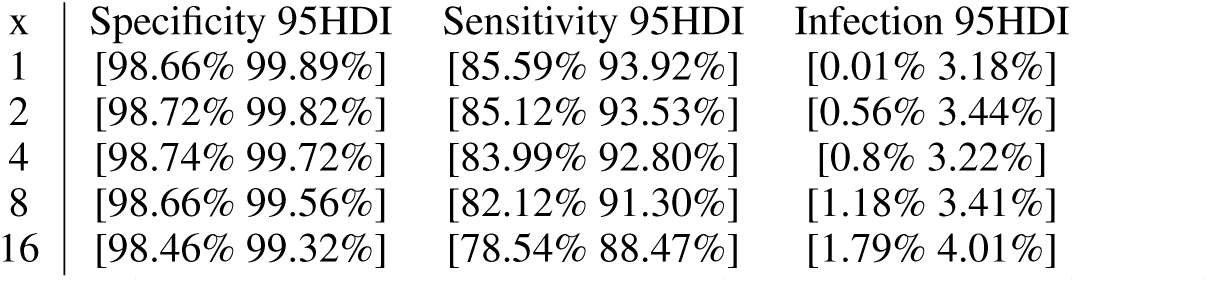
Sensitivity of our results based on our priors. 95% Highest density intervals for the variables of interest are presented for several possible priors.

